# ‘Finding light in the darkness’: exploring comedy as an intervention for eating disorder recovery

**DOI:** 10.1101/2023.04.18.23288749

**Authors:** Dieter Declercq, Eshika Kafle, Jade Peters, Sam Raby, Dave Chawner, James Blease, Una Foye

## Abstract

**Purpose:** Eating disorders remain a major health concern with the incidence of these illnesses has increased since the Covid-19 pandemic. With increased demands on treatments and service provision, and evidence that waiting for treatment is harmful, it is important that research explores novel and innovative adjuncts within interventions for individuals experiencing eating disorders. There is growing evidence highlighting that arts’ interventions are beneficial for recovery from eating disorders, with comedy workshops specifically, have shown promising results for recovery.

**Design:** The study used a qualitative interview study design, utilising pre, post and three-month follow-up individual interviews and workshop observations, to explore the feasibility of conducting a comedy intervention for people in recovery from eating disorders (n=10).

**Findings:** Findings show the comedy intervention had high acceptability and feasibility. For most individuals, participating in the course had positive outcomes, including promoting personal recovery across all five elements of the CHIME framework. Unique assets of the course included providing participants with an opportunity to distance themselves from everyday worries of living with an ED; the opportunity to cognitively reframe situations by making them the object of humour; and it provided a safe space to (re-)build a positive sense of self and develop self-acceptance through humour and play by challenging unhealthy perfectionism.

**Originality:** This study highlights that such innovative approaches can positively support personal recovery for people with eating disorders, with findings providing evidence that this comedy intervention acts to address elements of the CHIME framework to encourage and enable positive outcomes among participants.

---

Eating disorders (ED) remain a major health concern with a recent meta-analysis finding that the incidence of these illnesses has increased since the Covid-19 pandemic internationally, including in non-Western settings (*Silén & Keski-Rahkonen, 2022*). In the UK, there has been an increased demand for eating disorder services with a record demand for treatment being reported, with referrals record as increasing by almost two thirds since before the pandemic (*NHSE, 2022*). With such increasing incident rates and an increased demand for treatment, there is a need to focus on ways to support individuals’ recovery and treatment needs. Although there is guidance for ED patient care for providers only a minority of individuals affected by ED access effective evidence-based treatments (*Hudson et al., 2007*) particularly when compared to other psychiatric diagnoses (*Layard et al, 2012*). Research has found that many ED patients do not access evidence-based effective treatments, with estimates suggesting that in the UK only 23% of people with EDs receive treatment, a considerably smaller percentage when compared with 80% of those with psychosis (*Layard et al, 2012*). Furthermore, only a third of ED cases are detected by healthcare professionals (*Keski-Rahkonen et al, 2016)*. Given the evidence that waiting for treatment is harmful (*Furukawa et al, 2014*), it is important that research explores novel and innovative adjuncts within interventions for ED prevention and treatment to help and bridge this ever-growing gap between those needing care and those in services (*Nagata & Murray, 2021*).

Within the wider field of mental health, there have been calls for greater consideration to be given to alternative forms of therapeutic provision for people experiencing mental health difficulties *(Ostermann et al., 2019; Turgon et al., 2019*). Comedy workshops, specifically, have shown promising results for recovery from substance abuse, especially for psychological wellbeing, self-esteem, and self-efficacy (*Barker & Winship, 2016*). A recent review found that comedy and humour interventions are beneficial for mental health recovery and wellbeing and found preliminary evidence for a range of mechanisms through which comedy may have positive impact (*Kafle et al., 2022)*. The use of arts approaches has become increasingly popular within the field of eating disorders, with growing evidence highlighting that arts’ interventions are beneficial for recovery from eating disorders (*Ramsey-Wade & Devine., 2018; Smriti et al., 2022*).

## The Current Study: ‘Comedy for Coping’

Comedy for Coping (C4C) was created, developed and delivered by a stand-up comedian with lived experience of an eating disorder (DC). The workshop took place online (over Zoom) to minimize access barriers and accommodate Covid-19 restrictions. Workshop sessions ran for one hour, once a week, for six weeks (with a break of one week in the middle). The workshops train participants to deliver a stand-up comedy set to each other by the end of the series. Weekly sessions introduce key skills and understanding around attitude, stage presence, joke and set writing, and performance. In a typical week, participants would learn about comedy theory and participate in practical exercises. After the course was completed, participants received a weekly newsletter for 12 weeks, which included hints, tips and ideas on how to incorporate comedy into ongoing recovery.

While it is evident that there is preliminary evidence for the effectiveness of comedy interventions, the current picture created by research in this area, does not outline mechanisms of impact clearly. In line with the Medical Research Council guidance on evaluation and development of complex interventions, it is necessary that research should shed light upon which specific components of interventions have an impact on mental health (for specific people, in specific circumstances), rather than focus solely on whether interventions are effective (*Skivington et al., 2021)*. To unpack the ‘black box’ of comedy, there is a need for more studies that incorporate robust qualitative methods in their research design (*Kafle et al., 2022*). As a result, this study aims to explore the feasibility of conducting a comedy workshop intervention for people in recovery from eating disorders while understanding the core mechanisms of the course that may support personal recovery for participants.

## Study Methodology

### 4.1. Design

Qualitative interview study, using individual interviews and observations of the intervention’s sessions.

### 4.2. Research Team and Perspectives

This study was co-produced from inception, through design and in delivery by members with the comedian running the sessions (DC) and members of the research team including lived experience researchers (JP). Co-production work ran throughout the course of the study (e.g., via co-production of the study protocol, interview topic guide and analyses). Two researchers (UF & JP) led the interviews. Alongside members of the research team (DD & EK), the lived experience researcher (JP) conducted double coding of interview transcripts. This range of inputs was designed to increase the validity of our analyses; capturing content regarding aspects of recovery understood by experts by experience that may be lost when using a traditional researcher lead approach. Data analyses were jointly undertaken by all manuscript authors, with key themes established in an iterative process through review, reflection and discussion.

### 4.3. Inclusion and exclusion criteria

The inclusion criteria for participation were people who self-reported experience of an eating disorder, fluent in English and over 18 years old. People who were currently attending inpatient or day patient treatment (both NHS and private sector) were excluded, because the workshops are designed to support people in recovery (not in crisis). As the workshops took place online, people who didn’t have access to an internet connection and suitable device were also excluded.

### Sampling and Recruitment

Purposive sampling was used to ensure representation of a diverse range of participants. Information about the study was circulated via email and social media (e.g., Twitter) to a range of networks, including relevant professional organisations and networks, and workshops/conferences. Interested participants contacted our research team who provided further information about the study and ascertained whether they met the study inclusion criteria.

## Methods

To evaluate the impact of the workshops a range of methods were utilised to capture participants experiences.

### Interviews

Semi-structured interviews were conducted online in advance of the workshops, immediately after completion of the workshops, and after three months. This approach enabled us to understand participants recovery experiences, and their experiences of the workshops including if and how they support, or didn’t support, recovery. Semi-structured topic guides were developed by the research team to include questions which invited participants to reflect on their recovery, including explore recovery from the perspective of the CHIME framework. Interviews were conducted online via Microsoft Teams or Zoom during which participants read and completed online consent forms in advance of the interview, and the researcher reconfirmed consent at the start of the session with recorded verbal consent taken from all participants. Each interview lasted between 30 to 60Lminutes, with audio recordings interviews being transcribed by a member of the research team (JP). Transcripts for accuracy and all data was anonymised and allocated a unique ID number.

### Reflective Diaries

In addition to interviews, participants were asked to submit weekly reflective written diaries, which enables participants to voice their subjective and lived “reality” of recovery experience after each session during the workshop series (*Riessman 1993*).

### Observations

This method is increasingly used in health care research and allows for discrete observation of a community where participants have specific knowledge about an identified problem (*Higginbottom et al., 2013*). All workshop sessions were recorded (with the written consent of all participants attending) to allow a member of the research team (JP & EK) to observe the content of the workshops, the interactions around session activities, their engagement with activities, group dynamics, and the general running of the sessions. This recorded approach was designed as all workshops were conducted online and it was felt that a researcher being present would detract from the session. Observations were recorded as field notes and were uploaded into a Microsoft Excel spreadsheet.

### Data Analysis

We conducted a framework analysis (*Ritchie & Lewis, 2003*) informed by the CHIME framework of personal recovery. The CHIME framework was developed as means towards understanding the complexity of personal recovery which covers five components of effective recovery-oriented services and interventions including Connectedness, Hope, Identity, Meaning and Empowerment (*Leamy et al., 2011*). These components have been found to be prevalent across the research literature (*van Weeghel et al., 2019*), including with eating disorder populations (*Wetzler et al., 2020*). Given the lack of definitional cohesion and consensus in ED recovery (*Bardone-Cone, Hunt & Watson, 2018*), this study utilised the theoretical framework on personal recovery (PR). Differing from clinically based recovery, e.g., the absence of symptoms, PR recognises recovery as a journey towards a greater quality of life, achieved through adaptation and thriving despite mental ill health or ongoing symptoms (*Repper & Perkins, 2003; Davidson, Lawless & Leary, 2005; Leamy et al., 2011)*. Specific to EDs, literature suggests that patients view of recovery align with this PR conceptualisation (*Bohrer, Foye & Jewell, 2020; Pettersen and Rosenvinge, 2002*).

Our coding framework used the five elements of CHIME (*Leamy et al., 2011*) to record expectations and experiences of the intervention aligning to Connectedness, Hope, Identity, Meaning in Life and Empowerment using a deductive coding approach. To ensure that the analysis captured elements of recovery and experiences not aligned with the framework a secondary inductive coding was conducted by the team to pick up themes outside of the framework that may explore recovery and impacts that sit outside of the framework.

This initial stage was undertaken by two members of the study team with each researcher charting data summaries onto the framework for each of the interviews (EK, JP). Sub-themes within each broad deductive theme from our initial framework were then derived inductively through further coding and collaborative discussion within the research team (DD, UF).

### Ethics

Ethical approval was granted by the University of Kent Ethics Committee: Central Research Ethics Advisory Group. Ref: CREAG070-07-2021.

## Findings

### Section 1: about the intervention

#### Participant Characteristics

Participants were recruited via local and regional eating disorder charity networks, and via media promotion including social media posts and coverage in national and local radio, TV and print press (e.g., national newspapers). Of the 48 individuals who contacted the research team to take part in the study, a total of ten participants were accepted onto the course and attended the course. Selection of these ten participants was based on availability of the individual on the dates and times of the course, alongside an anonymised selection process to ensure a diverse range of individuals based on gender, age, ethnicity, diagnosis and geographical location.

Of the final participants, all had identified themselves as in recovery for a variety of eating disorders. Four people had Anorexia (restrictive); one person had Anorexia (binge-purge); one person had AFRID; two people had binge-eating disorder; and two people had Bulimia. Participants ages ranged from 25-46, and the majority (n=9) identified as being from a White background with one participant identifying as British Asian. The majority of the group identified as female (n=8) and two identified as males; one participant identified as transgender.

#### Attendance and Adherence, demand for the course

Overall attendance for the course was 83% with three of the fix weeks having 100% attendance (see table 1 for weekly attendance rates). Two participants dropped out of the course overall; one due to health reasons, while another was due to work related issues. Reasons for non-attendance from other participants included internet connectivity issues, covid-19 and scheduling challenges for childcare and work events.

**Table 1:**
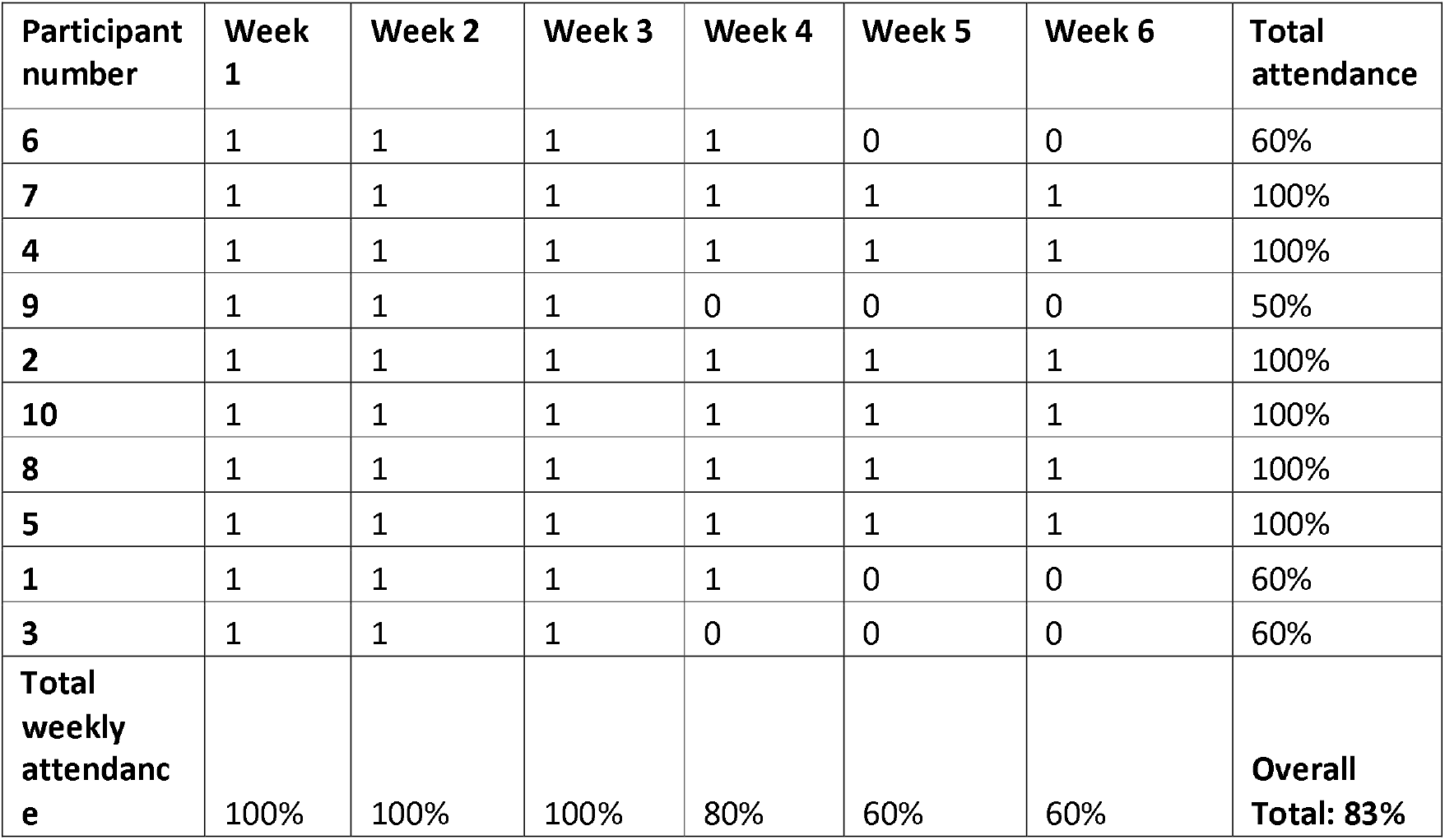
Participant attendance by week

**Table 2:** Themes organised by CHIME categories

| CHIME Category | Themes |
| --- | --- |
| 1. Connection | 1.1. A sense of community<br>1.2. Building connection through being part of an alternative peer support group<br>1.3. Connection beyond the group |
| 2. Hope and Optimism | 2.1. 'It's easier to be optimistic when you're laughing and having fun'<br>2.2. Positive role modelling recovery |
| 3. Identity | 3.1. (re)Building a sense of self: a space to really be me<br>3.2. Creating distance: 'I no longer want to be defined by my eating disorder' |
| 4. Meaning in Life | 4.1. Finding enjoyment in life: meaningful goals and roles<br>4.2. Accepting and normalising illness |
| 5. Empowerment | 5.1. 'Building yourself up': the importance of confidence and self-esteem<br>5.2. 'Taking responsibility for my own recovery' |

## Findings

Overall, participants reported a good level of acceptability of using comedy as part of an intervention for recovery for an eating disorder, irrespective of their ED diagnosis. Furthermore, the nature of the course made it highly acceptable to those who felt they did not fit the ‘*traditional eating disorders image’*, e.g., those who identified as male, from non-white backgrounds, and those with binge eating disorder. Using the CHIME framework, the findings from data collected before and after the course, as well as at a three-month follow-up period are categories below to exemplify the core mechanisms of the course that were felt by participants as salient within their recovery.

### 1. Connection

#### 1.1. A sense of community

Connectedness was highlighted by all participants as something that was valuable to their recovery but was challenging when whilst living with an eating disorder. Many cited this as an important reason for reaching out to attend the course.

> “*Eating disorders are very isolating illnesses, and I think it’s very common to feel like you’re the only person feeling a certain way*… *if it [my ED] gets really bad again, I will lose friends. Like I said, a huge part of this sort of disconnect is declining invitations to things to listen to your eating disorder”* (Participant 1, pre-course interview).

These expectations to support connections with others spanned participants inner network such as better relationships with family and friends, as well as with new people including with others who had similar experiences.

> “M*y instinct is always to be alone but I’d like to be more sociable*” (Participant 5, pre-course interview).

The course was seen as a way to connect with others who had similar experiences so participants wouldn’t feel so alone in their recovery journey, as well as being able to connect with others who understood the challenges and ups and downs of recovery and relapse. Despite having an eating disorder for many years, this course provided an opportunity for some participants to openly talk about their experiences for the first time;

> *“I guess you’re the first person I’ve spoken to who’s got an eating disorder?”* (Participant 4, pre-course interview).

This was particularly salient for those with experience of more stigmatised or unseen eating disorders, with one participant reporting that he had “*not ever spoken to anyone else before about binge eating disorder*” (Participant 6, pre-course interview).

Additionally, this participant highlighted specific gender-related challenges, explaining that it felt more challenging for him as he was male and eating disorders are often framed as a female issue. The fact the course was led by a man helped him feel more able to connect with others and to feel safer in this setting;

> “*I’ve got a friend, who had bulimia and he always talks about it, and so that was my first and only experience of another bloke talking about their eating disorder*” (Participant 6, pre-course interview).

After completing the course, participants reflected on the he shared experiences of the group as an important element that helped reduce their isolation and feelings of being alone in their recovery journey;

> *“I think if I learned anything, it’s that actually maybe I need to be in the company sometimes with other people who have eating disorders… I felt really comfortable and secure in that group of people in a way that I never experienced, really, in a group of total strangers. Because I always carry this sort of secret burden around with me all the time.”* (*Participant 7, post-course)*

> *“I enjoyed being with people who had the same weird thought processes as me and being together in a supportive way was helping” (participant 5, reflective diary, week 3)*.

From the first week of the course, a feeling of connection was felt by a number of participants with the shared experience of having an eating disorder and peer support emerging highlighted as a key mechanism building this sense of community;

> “*[a]lthough, really, we were just a group of strangers with eating disorders talking to one another on the internet, the task brought us together in a way I’d never expected… it really did feel as though we were all in it together”* (Participant 1, Reflective Diary, week 1).

This experience of connectedness and community during the meetings was felt after the course was completed and appeared to be sustained into the three-month follow-up;

> *“I think that’s what I got out of it the most was the sense of community… By the end of the sessions, we were all really good friends; I really wasn’t expecting it. And it was like an added bonus*” (Participant 8, post-course interview).

As Participant 8 continued to explore his engagement on the course, it became clear that connectedness felt strongly related to his recovery;

> *“It feels like its related… I think being in being in a space with other people with eating disorders because I know like two or three friends off the top of my head [made during the course]. I think that really helped. Yeah, like, I’m not sure how it helped but it clearly has because I’m a healthier weight and all round healthier in general. So it must have done (Participant 8, 3-months interview)*.

#### 1.2. Building connection through being part of an alternative peer support community

Participants reflected in the post-course interviews that the nature of the intervention as focused on comedy rather than the eating disorder was an important element of the group work. The majority of participants found the focus being placed on recovery rather than illness was key, and an important element as it allowed participants to connect across a range of experiences and diagnoses;

> “*I loved being in a self-help group that wasn’t talking about eating disorders (…). It was just such a lovely, everyone’s supportive, everyone understanding each other without ever talking about it”* (Participant 6, post-course interview).

For participants who had experienced traditional self-help groups, this approach was refreshing as it moved away from approaches that focus on taking away the eating disorder and instead was looking towards what recovery can give you back;

> *“I’ve done group support stuff before, [but] I never really gelled with it… it felt almost like just comparing notes rather than in any way talking about, like getting better. And although we didn’t talk about that, in the comedy, course, what I really liked was knowing everybody was experiencing similar difficulties… it made me feel better about the shame I feel. It was such a relief, to be in an environment where, everyone knew I had an eating disorder, and no one was asking me if I got better that day* (Participant 7, post-course).

Participants reported that they wanted the course to last longer to further elongate these connections and build from this, with requests post-course including a want to meet in person at follow-up stages, having spaces to contact everyone or having a newsletter to know how everyone is doing. This shows the real connections made within the group and the desire to continue fostering the positives from sharing this experience with others who understood their recovery journey. The use of newsletters that were circulated for 12 weeks following completion of the course, which included during the Christmas holiday period, were felt to help achieve this and was considered a positive addition to the course and refocus them on their recovery by reminding participant about these connection and the allyship that existed in the wider world supporting their recovery.

> “*I enjoyed receiving the tips and certain things to focus on that [the course leader] puts in the newsletter. That’s like, just remembering. Yeah, there are other people going through this too*” (Participant 1, 3-months interview).

#### 1.3. Connections beyond the Group

For several participants, the course also stimulated further connectedness in existing relationships through building friendships with others on the course that expanded beyond the sessions. This was salient for individuals as they had noted the negative impact on their core connections as they felt that they didn’t have the words or tools to do so due to the rigidity of their thinking related to their eating disorder. By doing the course, they noted that the comedy elements of the course acted to foster these connections with existing friends and acquaintances.

> *“I think sometimes I can kind of feel a bit like I don’t know what to start with talking about stuff that’s actually going on with me but I think [the course has] certainly gave me something to talk about with people, which is quite nice*. (Participant 2, post-course interview).

Comedy worked as a universal language for participants to share the experience of living with an eating disorder with their loved ones and helped participants to strengthen bonds with loved ones, and helped to remove the sense of shame and stigma around talking about eating disorders;

> *“Because my husband knew I was doing [the course], he would ask what are you doing? And I would never ever talk about my eating disorder ever with him, he knows, but we never talked about it. And it just made it a bit more normal. (…) I think we learned stuff about each other, which was really nice – we’ve been married for 11 years.”* (Participant 7, 3 months interview).

Similarly, one participant noted that he felt the skills from the course helped him connect with friends by making the topic of his eating disorder more approachable for friends thus opened up avenues to share and discuss when things related to recovery were difficult or frustrating.

> *“Since doing comedy for coping, everybody around me has learned that it’s okay to laugh with me about that sort of thing*… *it’s made it easier to speak about for myself and for people that I’m speaking about it with because it’s sort of like more accessible. I feel a bit better talking about that sort of serious stuff now*.” (Participant 8, post-course).

### 2. Hope and optimism: *finding the light in the dark of recovery*

#### 2.1. It’s easier to be optimistic when you’re laughing and having fun

The comedy element of the course was noted as an important component of the course as participants felt that it provided a light in the darkness of their journey, and thus reframed recovery as something fun and positive rather than something to fear. As a result, a few of the participants noted that this helped build a sense of hope and optimism about recovery therefore felt confident to make choices to engage in recovery in a hopeful manner:

> *“The course helped in how I could, you know, make quite a dark subject matter lighter*… *it helped me think about things differently and explore the subject matter in a safe and sort of controlled way. Particularly thinking about mental health… actually just reframing things it made the effect of them feel less negative.”* (Participant 6, post-course interview)

For one participant who continued performing stand-up comedy after the course ended, comedy became an active part of their recovery and a reason for them to continue to keep well thus actively choosing recovery.

> “*When things do go to shit I have started saying to myself, no, there’s gotta be some stand up material in this somewhere, which is a nicer way of thinking about the whole world exploding around me, but I bet I can make a joke somehow”* (Participant 2, post-course).

This positive stance and reframing approach was seen as unique when compared to other peer group or therapeutic experiences with the element of fun provided by comedy, allowing people to feel more enthused to choose recovery. This was contrasted to traditional therapies, which felt emotionally draining:

> *“It was like therapy, but in a really nice way because like when you have therapy quite often, you feel drained and like a bit sad afterwards (…) But after every, like, session of the course, I just felt like a little bit more alive and definitely a lot happier* (3-months interview).

##### 2.2. Positive role modelling recovery

Participants explained that the course leader, who has lived experience of an eating disorder functioned as a positive role model who embodied the optimistic perspective that recovery gives something back and can be fun.

> “*It must be something to do with [the course leader] I thought that he was able to be vulnerable, and funny and instantly put everybody at ease. (…) I think he established that. And then everyone just went with it. And once one person, like allows themselves to be vulnerable, it just builds*” (Participant 7, 3-months interview).

For Participant 6, the influence of the course leader as a role model further expanded to professional dreams and aspirations;

> “*I’ve learned so much from [the course leader] … it’s given me motivation, [the course leader] has done, so I can do it too… (…) he gave me the confidence to get stuck in and try stuff*” (Participant 6, post-course interview).

This also exemplified the importance of lived experience within the course as provided a role model who identified as recovered, showing that recovery is possible which was viewed as a necessary element of the course that instilled hope in participants to keep going.

> *“I also just generally find [the course leader] someone who makes me feel genuinely quite hopeful… he’s just a good role model. Having it delivered by somebody who openly will say that they’ve been there is and is no longer in that place is, yeah, that gave me a lot of hope*. (Participant 2, post-course interview)

#### 3. Identity: *I am more than my eating disorder*

##### 3.1. (re)Building a sense of self: a space to really be me

For several participants, the course offered a space where they could “be themselves”. The exercises gave permission to explore who they were in q playful, exploratory way. This was important for recovery as participants noted that they get rid of the eating disorder made them lose themselves and the course provided the safe space to explore these elements of identity that they may have lost or may not have considered as important to their recovery journey.

> *“[the course] offered a kind of solace I hadn’t been able to find anywhere else; a safe, supportive, uplifting space, where I felt comfortable enough to let go of everything that had been weighing me down, and really be myself… it’s a space for an hour a week where I could be me… it’s only when you do something that reignites that part of you that you realise it’s been missing*” (Participant 1, post-course).

For one participant, *s*he felt that the invitation to identify topics for making comedy as an opportunity to look “*at the multifaceted nature of yourself as an individual*” and that the course had provided not just an opportunity to rebuild identity but create a new sense of identity. This was particularly salient as recovery was felt to be about finding who you want to be not return to who you used to be before the eating disorder for this participant.

Doing “*something completely new really has been a good thing on the identity front of allowing me to kind of I don’t know, not rebuild, just create something new*” (Participant 2, 3-months interview).

##### 3.2. Creating distance: I no longer want to be defined by my eating disorder

As the course focused on recovery and didn’t place emphasis on the eating disorder, it presented a space where participants felt safe to be themselves and explore who they are. This was something that participants felt was difficult to do in their everyday life as loved ones would often focus on the eating disorder due to concern for them thus became a defining feature of who they are in that relationship. During the course, the shared experiences and lack of focus on these experiences allowed for a freedom to explore freely who they are and who they want to be away from the eating disorder.

> *“Yes, we were talking about mental health and mental illness and eating disorders, but not in a way that it defined who we were… I would always came away feeling that there was a part of me that had come alive for that time, and I realised in terms of identity and stuff, that’s the part that gets squashed by my eating disorder”* (Participant 1, post-course interview).

This change was sustained after the course for more than one participant who three months after the course noted they no longer felt defined by their eating disorder.

> *“I have a more solid sense of my identity now. (…) [I]f you asked me to write a list of all the things that make me me, I don’t think I’d even put my eating disorder on it. Because it wouldn’t occur to me*.” (Participant 7, 3-months course interview).

#### 4. Meaning in life

##### 4.1. Finding enjoyment in life: meaningful goals and roles

While the course was intended to support people with their eating disorder recovery, its impact spanned beyond this with a number of participants noting that the course gave them more tangible social goals and roles. The creative nature of the course allowed them the space to think about new hobbies, creative outlets and plans for the future, allowing participants to create new aspirations or goals for their future;

> “*I’d really like to write a book… and I’ve learned so much from [the course leader], that sort of humour side of it, I can now actually employ and weave that into my work. And it’s given me motivation*” (Participant 6, post-course interview).

For participant 2, stand-up comedy itself became socially and professionally significant as after the course completed, she explained she stayed in the habit of “*doing it [stand-up comedy] quite regularly. In fact, I’ve just got my first paid booking*” (Participant 2, post-course interview).

This was important for people’s recovery as it great their sense of meaning in life beyond an eating disorder and gave them a reason to keep engaging with recovery to allow them to engage in these new hobbies, interests and goals.

> *“Even if I have a bad day, now I get out of bed the next day, like with hope and optimism*. (Participant 7, 3-monhts interview).

For those earlier in their recovery journey noted that the course was able to remind them that there are things they enjoy, even in the depths of a bad day and served as a reminder that there is more to their life than the eating disorder.

> *“It has done me a lot of good to engage with a hobby… it really fits into the work I’m doing with my therapist who has said* ‘*you need to go and find something that, during this really hard time you’re having, is just fun*’*… the course did that, it boosted my spirits so when I was having a down day and its harder to engage in my meal plan can turn to the [courses skills] to make everything a bit easier.”* (Participant 2, post-course interview).

For participant 2, who continued with stand-up comedy, the benefits to self-management were particularly tangible after completing the course, as keeping well was important to be able to keep doing stand-up comedy:

> “*I’m aware, if I go back into fully eating disorder mode, I won’t be able to do it [stand-up comedy]*” (Participant 2, 3-monhts interview).

##### 4.1. Accepting and normalising their experience

Participants noted that the various elements of the course not only gave a positive spin to recovery but also by finding the light in these experiences, they were able to have a greater acceptance of their eating disorder to help push forward in recovery. Part of this included accepting that the eating disorder as part of their life but not the defining element of them.

> *I’m not allowing at the moment for my eating disorder to be like the primary thing in my life, I feel so much more accepting of it*” (Participant 7, 3-months course).

Linked to feelings of connection, participant reported that sharing space with other people with similar experiences and with the course being peerled with others helped for them to feel more normal in their journey thus helping them to accept that it isn’t that they are broken or disordered.

> *“Never in my life have I been in a room with someone outside of a therapy situation who’s talked about problems with food… the group context definitely helped with the normalisation of we’re all here for similar reasons and no one is weird… I don’t feel like some sort of alien anymore… I don’t feel broken anymore*.” (Participant 7, 3-monhts interview).

Furthermore, the social nature of comedy as something people easily connect with helped participants to normalise eating disorders and mental health problems within their inner circle and loved ones, therefore helped to make discussions about recovery as more accessible and acceptable.

> “*It’s easier to say to someone, I’m doing this stand-up thing, which is looking at recovery from eating disorders than it is to say, I’m doing CBT again, or I’m doing Mantra, or I’m seeing my dietician this morning*.” (Participant 2, post-course interview).

#### 5. Empowerment

##### 5.1. Building yourself up: the importance of confidence and self-esteem

Before the course, most participants felt nervous about being funny or confident. While there was a tension about whether theperf course was really for them, there was a sense of achievement for many for having attended and pushed themselves outside of their comfort zone.

> “*I’m so proud that I actually committed to something because normally, when it comes to anything relating to my issues, I’ve failed, I found fault or I’ve not been, you know, couldn’t be bothered. But I am very proud that I continued and that I completed it*” (Participant 5, post-course interview).

For several participants, taking part in the course gave them a sense of achievement and optimism about their own abilities as it helped to encourage participants to see being funny/comedy as a strength and participants felt valued within the group;

> “*I felt quite happy that I was quite funny a couple of times. I felt like I’d achieved something. It was really nice*” *(Participant 7, post-course)*.

A sense of achievement and confidence in themselves was important for most participants as they had noted before the course that they had low self-esteem and lacked sense of self, something common across eating disorders. Building this sense of achievement instilled a sense of hope for recovery and self-confidence that had very real implications in their lives. For participant 2, they noted that this confidence had a direct impact on their recovery as it supported them to sustain hope in recovery thus helped to avoid relapsing during and after the course.

> *“The course has given me the confidence to go and do other activities that I’ve kind of engaged with because of the course, [otherwise] I would have probably been in a bit of a state where it’s like, there’s no reason to keep pushing forward*.” (Participant 2, 3-months interview).

The specific nature of comedy as a playful intervention provided participants with a safe space to build their confidence to challenge eating disorder thoughts by allowing them to fail and rather than allow themselves to crumble. The course allowed them to reappraise making mistakes as being a failure and instead allowed participants to find the humour in these situations, and allowed participants to challenge self-critical thoughts;

> *“Usually, I am my own biggest bully but it definitely helped me be a little bit more playful and learn to let go of the idea that everything has to be perfect and planned… [now] I can be a bit more spontaneous and can be like more light-hearted, and doesn’t have to take everything seriously and doesn’t have to catastrophise or like, do really like black and white thinking* (Participant 1, 3-months interview).

Participants explained that beyond their recovery, taking part in the workshops had positive impacts further into their lives as it had helped them to develop confidence around public speaking, which was useful for their personal and professional lives.

> “*If I look back on how I operated at work in terms of my confidence, like it’s worlds apart, I feel so much more comfortable with who I am. And like even public speaking… my self-esteem is slowly, I’m slowly starting to accept that I’m ok. And this course has been part of that. I can live with myself as I am.”* (Participant 7 post-course interview).

##### 5.2. Taking responsibility for my own recovery

Committing to the course became an opportunity for participants to secure a sense of personal responsibility for their own recovery. Having a sense of ownership and choice was central to engagement in the course as it allowed participants to actively chose recovery, something that wasn’t always the case within previous experiences with treatment and support.

> *“I wanted to do it [the course]. I was doing it for myself, like it wasn’t like work. It wasn’t like for money. It wasn’t because someone else was telling me to. It wasn’t like NHS therapy that they were forcing me into*.” (Participant 1, 3-months interview).

Even the act of attending the course on days when it would have been easier to not attend was felt as a key indicator to participants that they were choosing recovery that night, something that acted as an example of recovery in action.

> *“In showing up that night, I wasn’t just showing up for other people, or out of a sense of obligation – I was doing it for myself. Which, I suppose, is what recovering from an eating disorder (or any mental illness) is all about”* (Participant 1, 3-months interview).

> “*Almost every single week, I sat there thinking, I’m not sure I want to do this, but I felt like I made a commitment and not just to myself, but to the group. I made a commitment and I wanted to stick to i.t*” (Participant 7, 3 months).

For one participant, they expressed that engaging with personal responsibility over their own recovery gave them a greater sense of control in life thus helped to create distance from patterns imposed by eating disorder and thus helped empower her to engage with building her identity and life outside of the illness.

> *“[the course] certainly made me feel like I was more in control of choosing what to do with my life… more in control of being able to shape my identity and empowered of being who I want to be rather than who everyone was telling me to be.”* (Participant 2, post-course).

##### Feasibility of running courses: limitations and challenges

While acceptability for the course was high among participants, key learning from participants highlighted obstacles for providing such interventions among those with eating disorders and the need for careful consideration when reviewing the feasibility of such courses.

Firstly, while it was acknowledged and communicated that the course was not designed as a stand- alone “solution” for eating disorder recovery and that it acts as a wrap-around to compliment traditional therapy and treatments, there was a need to manage expectations of the course. One participant reported feeling disappointed that the course didn’t meet their expectations of ‘*being cured*’ and ‘*rewiring how my brain works’* which flagged the need for additional screening prior to the course to ensure that such expectations are managed outside of the group setting. As this participant had a longer journey within their eating disorder having lived with anorexia for over twenty years with limited successes in treatment and therapy, this course felt like ‘*last ditch attempt’* at ‘*real recovery*’. While gains and positives were reported by this participant, the course was less acceptable for her, with other course members noting they picked up on some negativity as a result. Consequently, some participants reflected that the course may benefit from “*making sure that people are at more of a similar point in where they’re at, and in their outlook*” (Participant 2, post-course interview).

Secondly, while the course had strict rules to not engage in tip sharing or discussion of weight, there is a need for ongoing careful considerations and risk management of the content shared by participants. As reflection by some participants there were certain remarks made by another participant, which were viewed as “*a bit triggering, and difficult to listen to*” (Participant 1, post-course interview) and “*a bit much*” (Participant 2, post-course interview). While these were considered to have been managed by the course leader effectively, this flags the important need for expertise when facilitating such courses to ensure an understanding of the nuance of behaviours and cognitions that may be triggering and risky.

## Discussion

This paper demonstrates the acceptability and feasibility of using a comedy intervention to support personal recovery for individuals with eating disorders. For most individuals, participating in the course added to quality of life and well-being alongside promoting personal recovery across all five elements of the CHIME framework. Unique assets of the course included providing participants with an opportunity to distance themselves from everyday worries of living with an ED; the opportunity to cognitively reframe situations by making them the object of humour; and it provided a safe space to (re-)build a positive sense of self, develop self-acceptance through humour and play which also supported changes to challenge unhealthy perfectionism. Furthermore, participants highly valued connecting to others with EDs with the lack of clinical focus on ED symptoms and outcome measures being central as this approach provided space find their own identity and voice. For most participants, the atypical framing of the course was unambivalently celebrated as a welcome change from more traditional forms of peer support. This sense of community and connection was central to building someone up to engage with recovery as it tackles the challenge of recovery but also to allowed them to re-engage in a word outside of the eating disorder. Such social elements have been noted in user-led definitions of recovery as important (*Bohrer, Foye & Jewell, 2020; Richmond et al., 2020*).

These findings therefore provide evidence for the potential positive impact that innovative arts-based interventions may have for personal recovery for individuals with eating disorders, adding to the growing literature supporting the potential of arts within this field (*Bucharova, Mala, Kantor & Svobodova, 2020)*. This paper however adds to this field by highlighting the use of comedy and humour as a uniquely therapeutic element that provides a positive stance to reframe recovery in positive, enjoyable and empowering. Using innovative ways to engage people in new activities and having change from the normal approaches helps people want to choose recovery. Such positivity is salient and central to within personal recovery definitions as this journey requires positive subjective experiences of internal transformation (e.g., hope, meaning, healing, empowerment, and connection to other people) alongside positive external conditions (e.g., recovery-oriented services, positive environments of healing, and human rights agenda) (*Wetzler et al., 2020*; *Andresen, Oades, & Caputi, 2003; Jacobsen & Greeley, 2001; Reisner, 2005*). As a result, interventions that encourage and empower recovery to be giving something back rather than taking the eating disorder away provide a positive breeding ground for recovery as they are less coercive, clinical, and demanding.

While few studies apply constructs of a personal recovery model to EDs (*Dawson, Rhodes, &Touyz, 2014; Piot et al., 2019*), this study adds to evidence that eating disorders recovery can be categorised by CHIME and that personal recovery definitions for eating disorders recovery is highly relevant for this population.

Furthermore, our findings replicate the growing evidence that personal recovery and clinical recovery can be mutually facilitatory (*Dubreucq et al., 2022)* with elements of clinical recovery such as weight gain and improvements in quality of life being cited among the outcomes for participants in this study following a personal recovery approach. As a result, innovative and personal recovery-oriented models and interventions that utilise such holistic approaches, such as that presented in this study can been seen to compliment clinical recovery, can provide additional options for individuals on waiting lists, those finishing treatment/ not meeting clinical thresholds, or for those who have limited options in traditional treatment models, e.g., binge eating disorder.

## Limitations

As a pilot, there are clear limitations to this study. Most obviously, the group of participants was small. Although the study recruited about three times as many participants, there were no resources to run additional workshop series. Follow-up studies with larger cohorts, and, ideally, control groups, are required to investigate if the results of this study can be reiterated at scale – and to quantitatively analyse the impact of participating in stand-up comedy workshops as opposed to other forms of recovery activity.

Finally, the study did not engage in market research to test whether there a wider audience of people with ED would engage with stand-up comedy as part of their recovery journeys. Recruitment was stopped when over three times as many candidates were recruited than there were spaces on the workshop. Media coverage and the social media network of the workshop leader were significant contributors to recruitment. To test the C4C intervention at scale, future studies would need to develop strategies to reach a wider audience. In this respect, although participants had a variety of EDs, the cohort was predominately white and female. It is a clear that a more developed recruitment strategy is required to reach a more diverse audience in terms of ethnicity and gender.

## Conclusion

Overall, this study highlights that such innovative approaches can positively support personal recovery for people with eating disorders, with findings from this evaluation providing evidence that this comedy intervention acts to address elements of the CHIME framework to encourage and enable positive outcomes among participants. Unique to this study was the evidence for the intervention impacting on the CHIME elements of meaning and purpose, something that previous studies into comedy interventions for mental health lacked evidence for (*Kafle et al., 2022*).

## Data Availability

All data produced in the present study are available upon reasonable request to the authors

## Acknowledgements

We would like to acknowledge all participants who took part in the intervention for their time, honesty and engagement with the study.

## Notes

**Availability of Data:** The data that support the findings of this study are available on request from the corresponding author. The data are not publicly available due to privacy or ethical restrictions.

### Competing Interest Statement

This paper presents research funded by the British Academy

### Funding Statement

This paper presents research funded by the British Academy.

